# Regulation-Driven Variation in Utilization and Cost of B-Cell Depleting Therapies for Multiple Sclerosis: A Cross-National Study

**DOI:** 10.64898/2026.08.19.26360842

**Authors:** Nicholas S. Streicher, Thomas Angell

## Abstract

**Background:** Rituximab and ocrelizumab target the same CD20 receptor. Rituximab is off-patent and prescribed off-label; ocrelizumab is licensed and patent-protected. Whether the resulting differences in utilization and cost reflect clinical value or regulatory structure has not been examined.

**Objective:** To determine whether utilization and cost of B-cell depleting therapy across six health systems track regulatory approval status more closely than comparative effectiveness.

**Methods:** We examined rituximab and ocrelizumab utilization and cost in Sweden, France, Germany, the United Kingdom, Italy and the United States (2016–2024). Costs were drawn from published national sources on a consistent ex-factory basis. Utilization was registry-measured for Sweden, France and Germany, measured from national claims for the United States, and estimated from indirect data for the United Kingdom and Italy. Weighted annual costs per patient on B-cell depleting therapy were modeled by Monte Carlo simulation (10,000 iterations).

**Results:** Rituximab constituted the near-totality of B-cell depleting therapy in Sweden but 2.3% to 18.7% of use in the other five systems. Mean annual cost per patient ranged from $3,014 (Sweden) to $52,506 (United States), a 17-fold difference, with the four other European systems between $18,140 and $26,262. Adopting Sweden’s utilization pattern was associated with modeled five-year per-patient differences in drug acquisition cost of $76,000 to $248,000.

**Conclusion:** Utilization and cost align more closely with regulatory approval status than with available effectiveness data. International reference pricing acts on the price of the licensed agent but leaves intact the regulatory asymmetry that determines which agent is prescribed.

**Précis:** Rituximab’s share of anti-CD20 use in multiple sclerosis, and its cost, track national off-label reimbursement rules rather than comparative effectiveness, across six high-income health systems.

**Highlights:** What is already known. Rituximab and ocrelizumab deplete the same B-cell population, and a phase 3 randomized trial has now reported rituximab non-inferior to ocrelizumab on the primary radiological endpoint. Rituximab is off-patent and prescribed off-label; ocrelizumab is licensed and patent-protected. Whether the large international differences in which agent is used, and at what cost, reflect clinical value or regulatory structure had not been examined.

What this study adds. Across Sweden, France, Germany, the United Kingdom, Italy and the United States, rituximab’s share of anti-CD20 use ranged from 96% to 2.3% and mean annual cost per treated patient from $3,014 to $52,506. Because United States and Swedish ex-factory prices differ by the same 2.80-fold for both drugs, the cost gap separates exactly into a price component and a utilization component.

What it means for decision making. Utilization tracked national off-label reimbursement rules rather than comparative effectiveness or price. Adopting Sweden’s utilization pattern corresponds to modeled five-year drug-acquisition differences of $76,000 to $248,000 per patient. International reference pricing acts on the price of the licensed agent and leaves the regulatory asymmetry intact, so measures that change which agent may be reimbursed, such as treatment compendia and off-label outcome tracking, are the operative levers.

## Background

### Multiple Sclerosis and B-Cell Depleting Therapies

Multiple sclerosis (MS) is a chronic inflammatory disease of the central nervous system affecting over 2.8 million people worldwide.[1] Relapsing-remitting MS is the initial diagnosis in approximately 85% of people with MS,[21] and disease-modifying therapies (DMTs) are central to reducing relapse activity and disability accumulation.[2] Rituximab and ocrelizumab target the same CD20 receptor as off-label and on-label therapies respectively, and provide a uniquely informative comparison.[4]

Rituximab is an inexpensive chimeric monoclonal antibody whose patent protection has expired,[5] limiting commercial incentives to conduct the registration trials required for formal MS approval; licensing is therefore unlikely to be pursued. Off-label use is nonetheless widespread: the Atlas of MS reports rituximab as the most widely used high-efficacy disease-modifying therapy worldwide (78% of countries reporting, versus 75% for ocrelizumab), even though that ordering reverses with national income, ocrelizumab being reported in 90% of high-income countries against 80% for rituximab and in 92% of countries in the WHO European Region against 74%.[21] A 2025 Cochrane review of 28 studies and 37,443 participants concluded that, for preventing relapses in relapsing MS, rituximab as first-choice and as switching treatment compares favorably with a wide range of approved disease-modifying treatments.[66]

The financial implications are considerable. US direct healthcare costs per person with MS average approximately $70,000 annually,[9] and disease-modifying therapies are the single largest component at an estimated $57,202–$92,719 per user.[10] Small differences in regulatory positioning therefore translate into large differences in cumulative expenditure.

### Regulatory Frameworks Shaping Utilization and Cost

The coexistence of a low-cost, off-label biologic and a high-cost, on-label alternative targeting the same antigen provides a unique opportunity to assess how regulatory environments, rather than therapeutic value, drive prescribing behavior and healthcare expenditure. MS is particularly well-suited to this analysis as treatment decisions are long-term and DMTs dominate overall disease-related costs.[10,12]

We evaluated how regulatory frameworks across six healthcare systems shape the utilization and cost of rituximab and ocrelizumab in MS, comparing five large high-income economies with a conventional on-label/off-label distinction (the United States, Germany, the United Kingdom, France and Italy) against Sweden, where off-label rituximab is routinely used and reimbursed.

Together, these countries account for approximately 1.58 million people with MS (Atlas of MS, 2020),[21] yet represent highly varied approaches to pharmaceutical regulation in high-income settings.

Despite extensive discussion of drug pricing in MS,[11] the role of regulatory approval status in shaping cross-national utilization and costs of therapeutically similar B-cell depleting therapies remains unexamined.

## Methods

### Study Design and Data Sources

We conducted a comparative analysis of rituximab and ocrelizumab pricing and utilization for MS patients across six countries from 2016–2024. Utilization was defined as the proportion of patients receiving B-cell depleting therapy treated with a specific agent, except where noted (see Table 1 footnotes). Costs were expressed as mean annual per-patient drug acquisition expenditure.

**Table 1:**
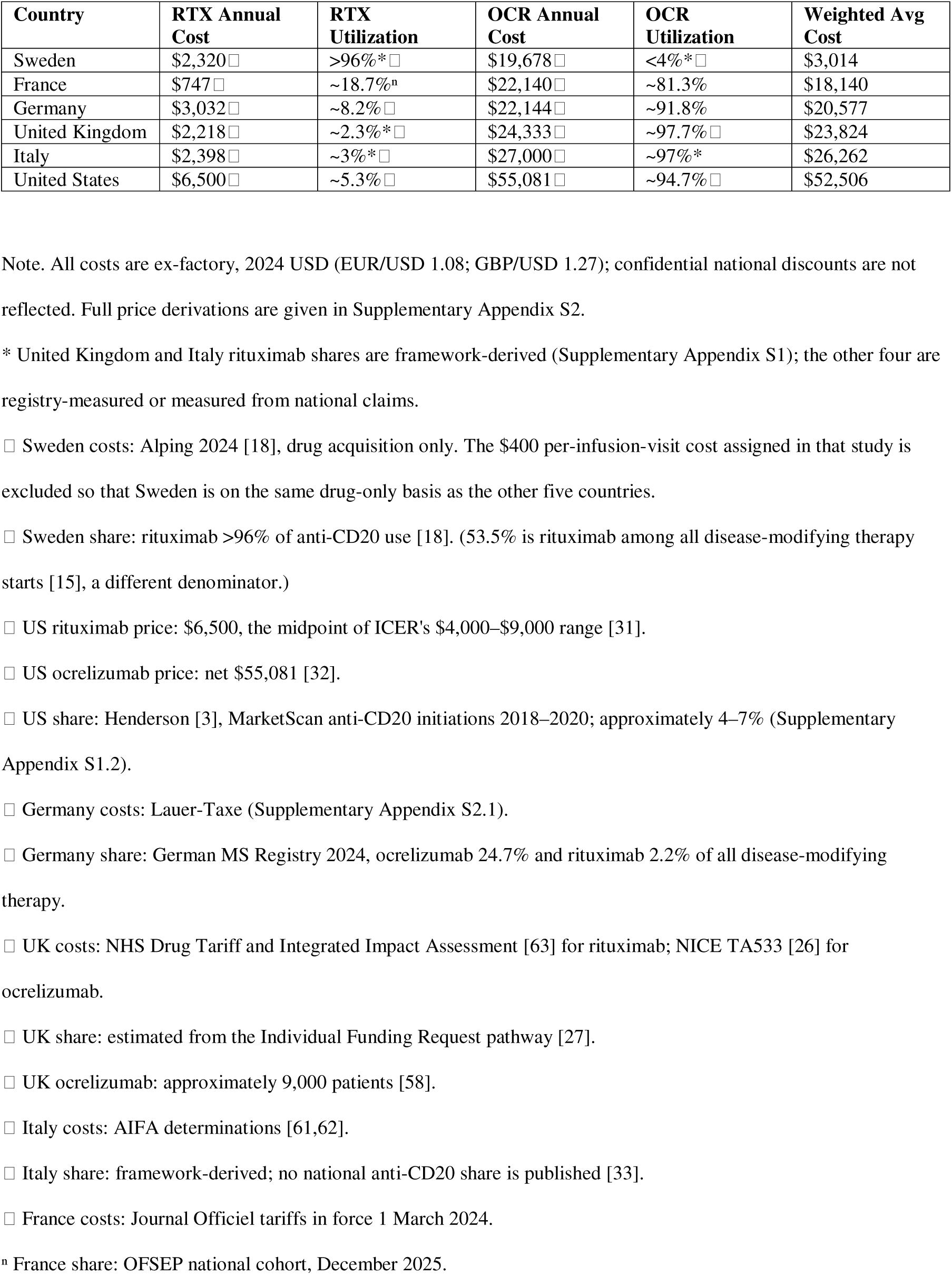
Comparative B-Cell Therapy Cost and Utilization in Multiple Sclerosis.

Data sources included the Swedish, German, French and Italian MS registries and cohorts,[14,22,30] national regulatory documents, and government price schedules (Journal Officiel, NHS Drug Tariff, Lauer-Taxe, Gazzetta Ufficiale).

All drug costs are reported on a consistent ex-factory (manufacturer) basis, excluding value-added tax and distribution margins, the basis used by the OECD, WHO and EURIPID for cross-national comparison. European figures are published list prices beneath which confidential discounts sit undisclosed; no discount percentage has been assumed or imputed. The United States is the single exception, where ICER publishes an explicit net-of-rebate figure, so comparing European list prices with a US net price is conservative with respect to our findings. Costs are expressed in 2024 US dollars (EUR/USD 1.08; GBP/USD 1.27). Full derivations, including the reverse application of the German Arzneimittelpreisverordnung, are given in the Supplementary Appendix (S2). National utilization data were unavailable for the United Kingdom and Italy. For these countries, utilization was derived using multiple estimation approaches informed by regulatory framework analysis and available regional or multicenter studies, modeled through Monte Carlo simulation as detailed in the Supplementary Appendix (S1, S5).

### Statistical Analysis

Monte Carlo simulations (10,000 iterations) propagated input uncertainty. Prices were drawn for all six countries using coefficients of variation of 0.083 to 0.14, and utilization from beta distributions for the two countries whose rituximab share is a framework estimate (the United Kingdom and Italy). Each iteration recomputed the weighted annual cost per patient receiving B-cell depleting therapy.

No null-hypothesis significance tests were applied to the simulated output: interval precision depends on iteration count rather than on any patient sample, so a p value on such output carries no inferential meaning. Each country’s mean weighted cost is reported with a 95% credible interval defined by its 2.5th and 97.5th percentiles, with cost ratios relative to Sweden and their corresponding intervals.

## Results

### National Utilization Patterns

Utilization varied markedly and paralleled national regulatory frameworks (Tables 1–2, Figure 1).

**Figure 1:**
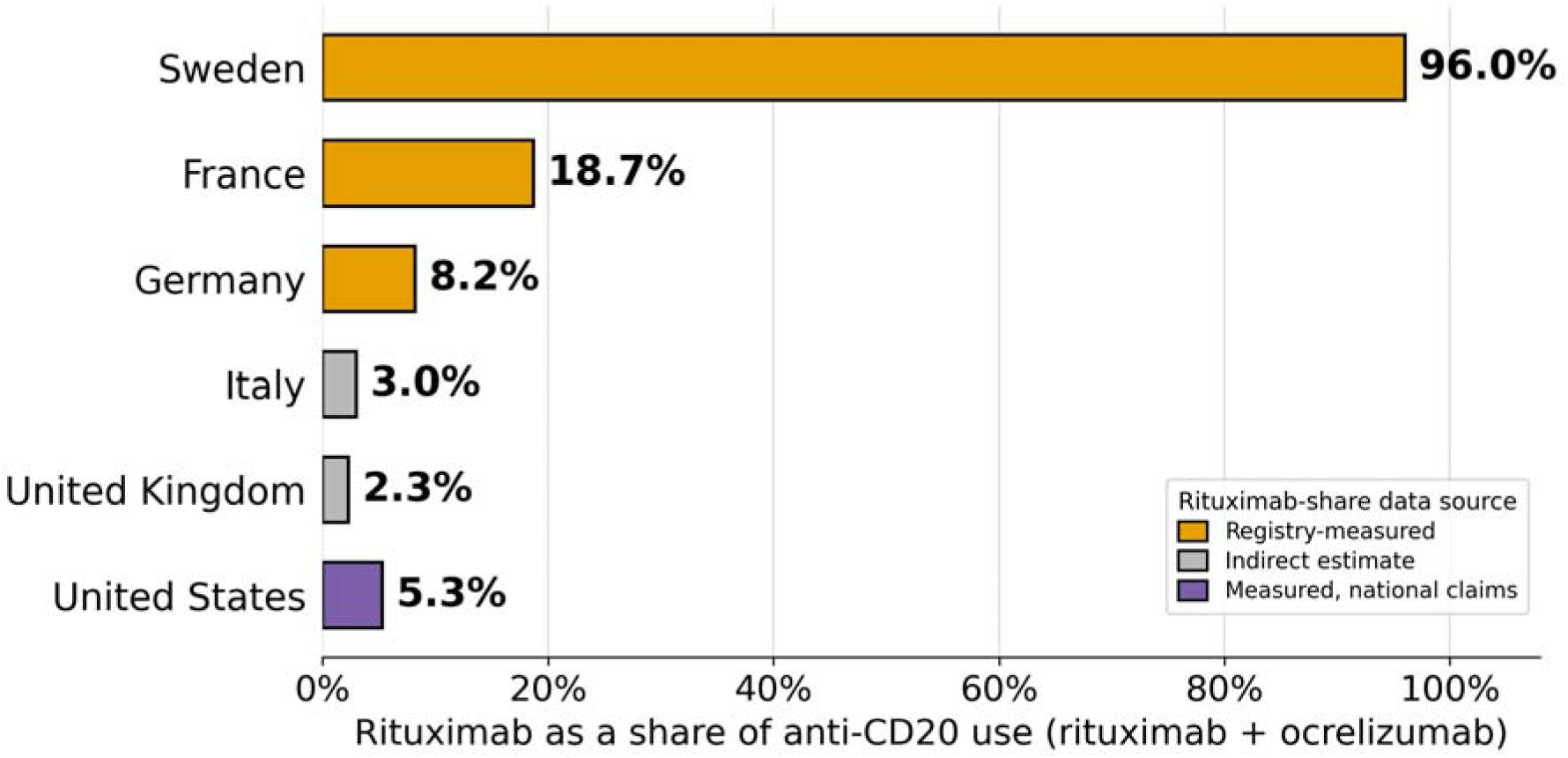
Rituximab Utilization Across Six Healthcare Systems. Rituximab as a share of anti-CD20 use (rituximab plus ocrelizumab) in each country. Bar color indicates the provenance of the share: registry-measured (amber), measured from national commercial claims (purple), or estimated from indirect data (grey). Sources and derivations for every country are given in Supplementary Appendix S1.

**Table 2:** Regulatory Frameworks, Pricing Mechanisms, and Utilization Data Quality by Country.

| Country | Off-Label Framework | Key Regulatory Barrier | Price Mechanism | Utilization data: coverage and limitations |
| --- | --- | --- | --- | --- |
| Sweden | Permitted; fully reimbursed | None; evidence-based prescribing norm | Regional negotiation | Swedish MS Registry, 20,678 patients at the end of 2024, approximately 87% coverage; represented at all 62 units providing specialised MS care [59]. Off-label rituximab is reimbursed and recorded on the same basis as licensed agents. The ocrelizumab share is not published and is inferred (Appendix S1.1). |
| Germany | Sandoglobulin ruling (2002) | No reimbursement if approved alt. exists | AMNOG negotiation | German MS Registry, more than 80,000 patients enrolled since 2001, about 29% of an estimated 280,000 nationally [60]. Rituximab is not broken out in headline reporting; participation by specialist centers is voluntary. |
| United Kingdom | IFR process required | Clinical exceptionality requirement | NICE negotiation | UK MS Register, more than 42,000 patients enrolled since 2011, about 31% of an estimated 134,000 nationally [60]. Off-label use is not captured, so the share is a structural low estimate. |
| United States | Permitted; coverage varies | Payer restrictions; formulary barriers | Market-based; rebates | No national registry; NARCOMS covers about 5% of an estimated 914,000 [60]. The share is measured from commercial claims initiations [3] and excludes public payers, patients aged 65 and over, and prevalent use. |
| Italy | Law 648/96 | AIFA list exclusion when alt. exists | AIFA negotiation | No national reporting of anti-CD20 utilization. The share is framework-derived from regional cohorts and Law 648/96, and is the least supported figure in this analysis. |
| France | Accès compassionnel (CSP L.5121-12-1); no compassionate-prescribing framework for MS | No appropriate authorized alternative | Tarif unifié (single tariff, originator and biosimilars); liste en sus | OFSEP national cohort, current-therapy export. Prevalent rather than initiation denominator; the share varies markedly by disease phenotype. |

*Note: Regulatory frameworks are described in Supplementary Appendix S3 and utilization sources in S1. Registry*
*coverage figures for Germany, the United Kingdom and the United States are from Stahmann et al. [60].*

Swedish practice permits off-label prescribing where the decision rests on scientific evidence and proven experience, and rituximab is fully reimbursed in this setting.[18] The Swedish MS Registry’s 2024 annual report gives current national figures: of 12,738 patients receiving disease-modifying therapy at the end of 2023, more than 7,400 were treated with rituximab, 58% to 63% of all such therapy, and 72.5% were receiving high-efficacy therapy with anti-CD20 agents or natalizumab. The registry describes rituximab as having become “completely dominant” in Swedish MS care despite lacking approval for the indication.[59] Ocrelizumab uptake is too low to include in comparative analyses,[18] so rituximab constitutes the near-totality of B-cell depleting therapy. On a drug-acquisition basis annual costs are approximately $2,320 for rituximab (500 mg twice yearly at $1,160 per 500 mg) and $19,678 for ocrelizumab ($9,839 per 600-mg dose twice yearly); the $400 per-infusion-visit cost assigned in the source study is excluded so that Sweden is reported on the same drug-only basis as the other five countries.[18]

The US demonstrates a contrasting pattern. Henderson et al. analyzed 153,846 disease-modifying therapy initiation episodes among commercially insured adults aged 18 to 64 in the MarketScan Commercial Claims and Encounters database; across 2018 to 2020, the years in which both anti-CD20 agents were established in practice, rituximab accounted for 5.3% of anti-CD20 initiations against 94.7% for ocrelizumab.[3] A second national claims cohort, drawn from Optum Clinformatics, of which 41.2% were enrolled in Medicare Advantage plans, gives a higher share of 17.3% [69], but accrued incident patients from 2014 and so predates ocrelizumab’s 2017 approval across more than half its span; pooling the two national series gives 7.4%, carried here as a sensitivity analysis.

Estimates from individual United States health systems diverge from these national figures (49.1% in an all-payer Colorado cohort [70]) for reasons the authors of that study attribute to longstanding local prescribing practice and to Medicaid access to off-label rituximab rather than to national practice. Every available United States figure is a share of treatment initiations rather than of prevalent patients, and therefore understates the prevalent rituximab share by an amount the published record does not identify. The sources, the composite estimate and the direction of bias in each are set out in the Supplementary Appendix (S1.2). Ocrelizumab is carried throughout at an annual acquisition cost of $55,081, a net-of-rebate model input reflecting a 23% discount to wholesale acquisition cost,[32] taken from the ICER 2023 report, which excluded rituximab from its base-case monoclonal antibody market basket despite documented off-label use and placed the price premium between rituximab and ocrelizumab’s net price at between 600% and 1,300%.[31]

Germany’s MS registry has enrolled more than 80,000 patients since 2001, approximately 29% of an estimated 280,000 nationally, with roughly 33,000 in its current documented cohort,[22,60] but does not break rituximab out in its headline figures. Its 2024 report gives the distribution of current immunomodulatory therapy among treated patients (n=8,733): ocrelizumab 24.7% and rituximab 2.2% of all disease-modifying therapy, so rituximab represents 8.2% of anti-CD20 use. Initiation data agree: in the same registry (n=2,658) rituximab accounted for 0.3% of first disease-modifying therapy starts against 3.5% for ocrelizumab.[60] Although the German Neurological Society’s 2021 guidelines group rituximab and ocrelizumab in the same efficacy category,[23,24] rituximab remains a small minority of use despite its lower annual cost ($3,032 vs $22,144), consistent with the 2002 Federal Social Court ruling in the Sandoglobulin case, which permits statutory health insurance to fund an off-label medicine only where the condition is serious, no approved alternative is available, and evidence supports a reasonable prospect of benefit.[25]

France occupies an intermediate position. Its statutory framework closely resembles Germany’s. Off-label prescribing under Article L.5121-12-1 of the Code de la santé publique is conditioned on the absence of an appropriate authorized treatment (the same test the Bundessozialgericht applies in Germany and that governs Italy’s Law 648/96 list), and the ANSM’s compassionate-prescribing framework for rituximab covers severe refractory immune thrombocytopenic purpura, not multiple sclerosis.[67,68] Nevertheless the OFSEP national cohort records rituximab in 1,431 of 7,646 patients receiving an anti-CD20 agent, a share of 18.7%, more than twice Germany’s and the highest of the four non-Swedish European systems studied. Two features distinguish France. Rituximab use concentrates where the statutory test is easiest to satisfy: 22.6% of primary progressive and 9.5% of secondary progressive patients receive it, against 1.2% of relapsing-remitting patients, for whom ocrelizumab carries a licensed indication. French rituximab is also the cheapest in the series at $747 per patient-year, a consequence of the tarif unifié, which fixes a single reimbursement tariff of €345.93 per 500 mg vial applying identically to the originator and to every biosimilar. France therefore returns the lowest weighted annual cost of the five non-Swedish systems, $18,140.

The United Kingdom presents a further pattern. Ocrelizumab was recommended by NICE for relapsing MS in 2018 and is routinely commissioned at a list price of £4,790 per 300 mg vial, giving £19,160 ($24,333) annually at the licensed dose, with an undisclosed patient access scheme applying.[26] Rituximab is not licensed for MS and is reachable only through Individual Funding Requests, 71% of which were declined in a tertiary-center series, most commonly for failure to demonstrate clinical exceptionality.[27,28] NHS England reported approximately 9,000 patients in England receiving ocrelizumab by infusion in July 2024.[58] No national dataset reports rituximab use in MS, so we estimate utilization at approximately 2.3% of B-cell depleting therapy recipients from this restrictive access pathway. Measured initiation data are consistent: in the UK MS Register (n=1,147) rituximab accounted for 0.1% of first disease-modifying therapy initiations against 3.0% for ocrelizumab,[60] and NICE states rituximab is not routine practice in the UK.[26]

The Italian MS Register[30] does not report aggregate rituximab utilization; Zecca et al. documented 355 rituximab-treated patients across 22 Italian and 1 Swiss center between 2013 and 2016.[33] Law 648/96 restricts off-label reimbursement when approved alternatives exist,[35] and the framework-derived estimate is 3%.

### Economic Impact

Monte Carlo simulation quantified economic implications across systems (Table 1, Figure 2). Mean annual per-patient costs were $3,014 (95% CrI $2,650–$3,418) in Sweden, $18,140 ($14,872–$21,997) in France, $20,577 ($16,858–$24,865) in Germany, $23,824 ($18,661–$29,861) in the United Kingdom, $26,262 ($20,696–$33,131) in Italy and $52,506 ($39,737–$68,209) in the United States. Sweden’s credible interval does not overlap that of any other country; cost ratios relative to Sweden were 6.0 (4.7–7.6) for France, 6.9 (5.4–8.6) for Germany, 7.9 (6.0–10.2) for the United Kingdom, 8.8 (6.6–11.3) for Italy and 17.5 (12.8–23.3) for the United States. All weighted averages use rituximab and ocrelizumab shares of B-cell depleting therapy as weights, applied uniformly across the six countries. For Sweden this differs from rituximab’s share of all disease-modifying therapy starts (53.5%),[15] since the remaining Swedish patients receive non-B-cell agents rather than ocrelizumab.

**Figure 2:**
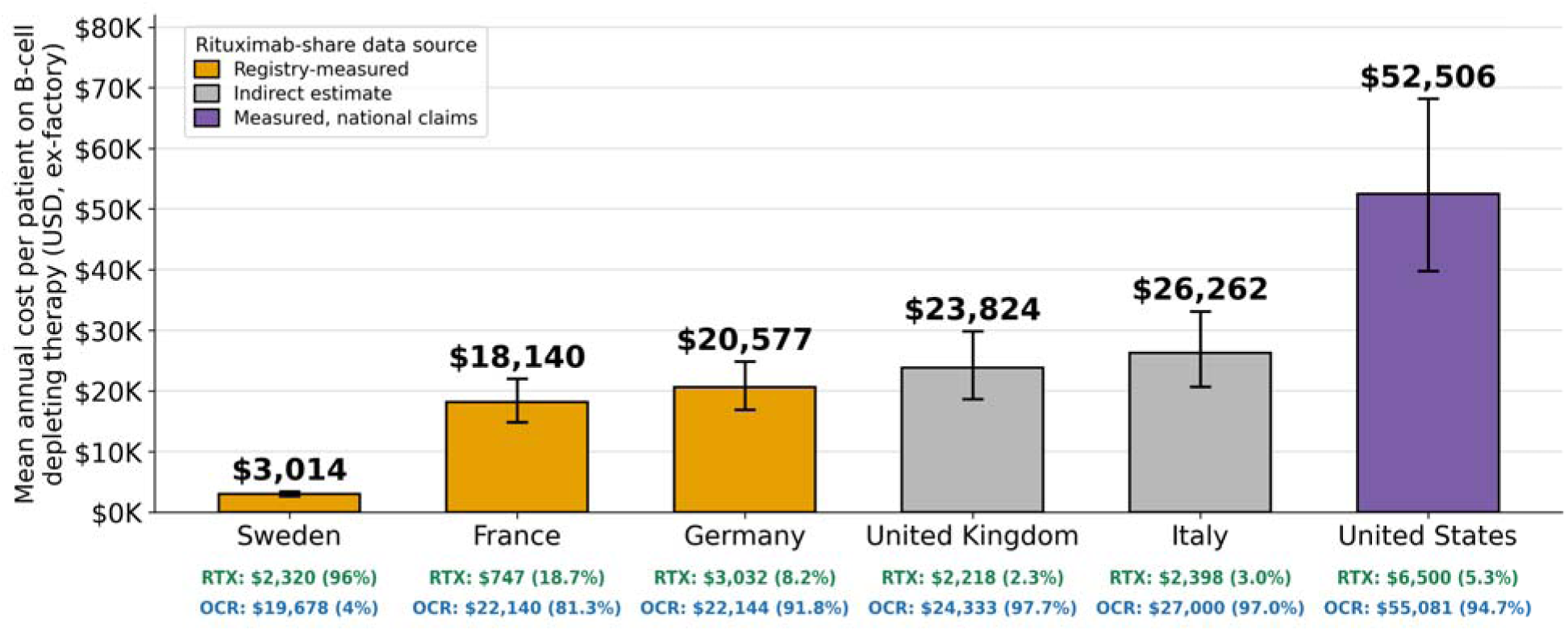
Mean Annual Per-Patient B-Cell Therapy Costs by Country. Mean annual cost per patient receiving B-cell depleting therapy, by country, on a consistent ex-factory basis. Bars are weighted averages from Monte Carlo simulation (10,000 iterations); error bars are 95% credible intervals. Bar color indicates the provenance of the rituximab share, as in Figure 1. Below each bar: per-drug annual cost and its share of B-cell depleting therapy. Price sources and derivations are given in Supplementary Appendix S2.

A Swedish nationwide cohort study reported total healthcare cost savings of $35,000–$66,000 per started therapy over five years for rituximab relative to natalizumab, fingolimod and dimethyl fumarate.[18] On the cross-national drug-acquisition basis modeled here, the gap between Sweden’s utilization pattern and those of the comparator systems corresponds to $76,000 (versus France) to $248,000 (versus the United States) per patient over five years. The magnitude of the aggregate opportunity depends on how many patients receive B-cell depleting therapy. As an exploratory illustration, if the approximately 5–7% of the roughly 1.58 million people with MS across the six countries who are treated with B-cell depleting therapy (on the order of 80,000–110,000 patients) achieved Sweden’s rituximab-forward utilization pattern, the resulting five-year difference in drug acquisition cost would be on the order of $13–$18 billion. This aggregate figure is a scenario estimate rather than a precise projection, as it depends on treatment-eligible population sizes that are not uniformly reported across countries; the per-patient differences, by contrast, follow directly from the country-specific prices and utilization shares in Table 1.

## Discussion

Between-country variation in weighted annual cost exceeded the variation in rituximab acquisition prices, so utilization rather than price alone was the principal determinant of expenditure. We found marked international variation in the utilization and cost of B-cell depleting therapies despite broadly similar comparative-effectiveness evidence. Across six healthcare systems, rituximab utilization consistently paralleled national regulatory and reimbursement frameworks rather than acquisition cost or currently available clinical evidence. These findings suggest that regulatory approval status functions as an independent determinant of treatment utilization once therapeutically comparable alternatives become available.

### Market Dynamics and Innovation Incentives

Ocrelizumab, ofatumumab and ublituximab all target CD20 and all remain patent-protected; rituximab does not. In US Medicare and Medicaid data ocrelizumab was priced 6- to 8-fold higher than rituximab ($69,949 vs $11,759 in Medicare, $47,671 vs $5,893 in Medicaid, fourth quarter 2021); at parity over 2018–2021 Medicare would have saved $1.9 billion, or 76% of its ocrelizumab spending, and Medicaid $590 million.[8] Rituximab appears on the World Health Organization’s Model List of Essential Medicines, which identifies it as a therapy that should be available at a price the health system can afford. The off-label agent is formally designated essential while its higher-priced on-label counterparts dominate utilization.

Rituximab demonstrated efficacy in the phase 2 HERMES trial, reducing gadolinium-enhancing lesions by 91% relative to placebo,[2] but ocrelizumab subsequently underwent the regulatory development required for approval while rituximab remained off-label.

Phase 3 trials cost $11.5–$52.9 million depending on therapeutic area,[42] revealing a paradox in pharmaceutical innovation: the system that funds clinical research by rewarding new indications simultaneously provides little incentive to test off-patent medications, which may disadvantage cost-effective therapies.

### Comparative Clinical Effectiveness and Safety

Until recently, head-to-head comparisons between B-cell depleting therapies were limited to observational cohorts. The OVERLORD-MS trial has now provided Level-1 evidence: in a phase 3, double-blind, noninferiority randomized controlled trial in newly diagnosed relapsing MS (218 randomized, 216 treated), rituximab met noninferiority to ocrelizumab on the primary MRI endpoint (absence of new or enlarging T2 lesions, 92.2% vs 94.8%; risk difference −2.6 percentage points, 95% CI −9.4 to 4.3; P=0.03 for noninferiority) over 24 months.[51] Secondary outcomes were closely comparable: annualized relapse rates were 0.09 (rituximab) versus 0.04 (ocrelizumab), and six-month confirmed disability progression was numerically lower with rituximab (3% vs 7%). This is consistent with prior observational data. In a Swedish cohort of 494 patients initiating a first disease-modifying therapy, rituximab showed lower discontinuation (annual rate 0.03 vs 0.29–0.53) and lower relapse and radiological activity than injectable therapies and dimethyl fumarate.[19] In RIFUND-MS, rituximab reduced the relative risk of relapse by 81% versus dimethyl fumarate (3% vs 16% over 24 months; risk ratio 0.19, 95% CI 0.06–0.62).[17] A prospective individual-patient-data meta-analysis pooling four randomized trials is expected in 2028.[36] The evidence is not uniformly favorable: Roos et al., matching 710 ocrelizumab- to 186 rituximab-treated patients, found rituximab did not meet noninferiority for relapse control (0.20 vs 0.09; rate ratio 1.8, 95% CI 1.4–2.4), though disability accumulation did not differ,[7] while a Finnish matched cohort found comparable effectiveness (annualized relapse rate 0.03 in both arms).[52] The randomized evidence supports noninferiority on the primary radiological endpoint and comparable disability outcomes, with some observational data suggesting a modest relapse-rate advantage for ocrelizumab. The residual difference is small relative to the price differential.

Swedish registry data further support rituximab: over five years it was associated with 0.12–0.22 fewer relapses per started therapy than each MS-approved comparator, and was dominant on both cost and relapse outcomes in that analysis.[18]

The safety comparison is balanced. In OVERLORD-MS, overall infections were more frequent with rituximab than ocrelizumab (82% vs 69%), driven largely by mild upper respiratory tract infections; serious adverse events were similar (8% vs 7%) and serious infections numerically identical at four participants in each arm.[51] The only cases of hypogammaglobulinemia and the only malignancy occurred in the ocrelizumab group, infusion-related reactions were comparable (23% vs 25%), and there were no deaths or opportunistic infections in either arm. Pharmacovigilance data are consistent: FDA Adverse Event Reporting System analysis found infections reported more frequently for ocrelizumab than rituximab (21.93% vs 11.05% of reports),[38] although these are proportions of spontaneous reports rather than incidence rates. Both agents carry the same rare but serious risks, progressive multifocal leukoencephalopathy[45] and hypogammaglobulinemia with prolonged use.[46] A nationwide Swedish cohort of 4,694 rituximab-treated patients reported serious infection at approximately twice the rate of comparator therapies (22.7 vs 10.4 per 1,000 person-years), a signal that did not vary by treatment line, prior therapy or exposure time.[20] On balance the randomized evidence shows a modest excess of largely non-serious infections with rituximab against equal serious-infection rates and a numerically more favorable profile for malignancy and hypogammaglobulinemia.

Rituximab and ocrelizumab differ in molecular structure: rituximab is a chimeric antibody (containing murine-derived sequences), while ocrelizumab is fully humanized, which theoretically reduces immunogenicity.[4] However, current clinical evidence does not demonstrate that this structural difference translates into meaningful differences in long-term efficacy, tolerability, or safety in MS treatment.

### Data Transparency and Policy Implications

Data availability varies systematically. Sweden’s registry publicly reports utilization,[14] Germany’s does not report rituximab in its headline figures despite guideline endorsement,[22,24] Italy’s covers approximately 50,000 patients without reporting off-label use,[30] and UK national datasets do not capture off-label prescribing. Without systematic tracking of off-label utilization, systems have limited capacity to assess whether higher costs are commensurate with clinical value.

### Potential Policy Solutions

Oncology offers a statutory model for integrating off-label evidence into coverage decisions. Under the Social Security Act, Medicare must treat an off-label use as a medically accepted indication where it is supported by one of the compendia recognized by the Centers for Medicare & Medicaid Services: AHFS Drug Information, Micromedex DrugDex, Clinical Pharmacology, Lexi-Drugs and the National Comprehensive Cancer Network (NCCN) Drugs and Biologics Compendium. Private insurers commonly follow. That pathway is confined by statute to anticancer chemotherapeutic regimens; no equivalent mechanism exists in neurology.[41]

To support this decision making, neurology could benefit from similar infrastructure to the Swedish MS registry that documents rituximab’s effectiveness in more than 4,900 MS patients[15,18]. Establishing neurology treatment compendia for systematic outcomes tracking could provide a standardized evidentiary basis for coverage decisions in MS as well as other conditions where there is documented clinical utility for off-label biologics.[48,49]

Physician cost awareness is limited, with fewer than one-third of drug-cost estimates falling within 20–25% of the true cost,[40] and real-time cost transparency at the point of prescribing reduced selection of high-cost agents with equally effective alternatives by 32%.[39]

Neurology compendia would require sustained funding, multistakeholder consensus and ongoing evidence review, though the cost disparities documented here suggest the savings may offset that investment.

These findings acquire additional salience amid a shifting international pricing landscape. In May 2025, US Executive Order 14297 established a “Most-Favored-Nation” (MFN) framework seeking to align US prescription drug prices with the lowest price paid by comparably developed nations, implemented through international reference pricing models and voluntary manufacturer agreements.[53] The cross-national gradient documented here, in which US ocrelizumab costs roughly 2.5-fold those in France, is the differential such policies target. Its consequences may not be confined to US prices.

Because reference pricing benchmarks US prices against foreign markets, manufacturers face a disincentive to launch or maintain products in lower-priced reference countries, and early analyses report falling European launches and rising withdrawals since the announcement.[54] Analysts have cautioned that convergence may occur upward rather than downward, consistent with the recently finalized US–UK arrangement, which raises UK net drug spending and the cost-effectiveness threshold.[55] These developments are early and causal attribution to MFN is not established. Genentech was among nine manufacturers entering such agreements in December 2025, each extending most-favored-nation prices to state Medicaid programs, discounting direct-to-consumer sales, and guaranteeing most-favored-nation pricing on new launches in exchange for tariff relief.[56,65] No reduction has been announced for ocrelizumab, and the only multiple sclerosis therapy named to date is siponimod. The principal consumer-facing mechanism, discounted direct-to-consumer sales, expressly excludes injectable and infused medicines, and the agreements do not require reductions in the list prices of existing products.[56] Infused anti-CD20 monoclonal antibodies, the drug class examined here, therefore sit largely outside the mechanism by which these policies are expected to lower prices.

Should branded prices converge across jurisdictions while access to newly launched therapies narrows in Europe, the case for an inexpensive, off-patent, randomized-trial-validated alternative would be strengthened rather than diminished. The regulatory barriers that limit its use would become a more consequential determinant of what patients receive.

### Limitations

National utilization data were available from Sweden, Germany, France and the United States, the last measured from national commercial claims.[3] For the United Kingdom and Italy, we relied on regulatory framework analysis, as comprehensive national statistics remain unreported. Confidential discounts beneath the published prices are not disclosed and were not imputed. Swedish all-DMT initiation data (53.5%) date from 2018 and may not reflect current patterns; the model input is the 96% anti-CD20 share.

Because the European figures are list (ex-factory) prices with confidential national discounts beneath them, they are ceilings rather than realized costs, whereas the US ocrelizumab figure is net of rebates; this asymmetry is conservative with respect to our findings. In Germany, office-based ocrelizumab dispensed at the community-pharmacy retail price would raise total system outlay above the manufacturer price reported here. Finally, a single all-forms share per country mixes a relapsing population, where the head-to-head evidence is strongest, with progressive patients treated with rituximab because alternatives are scarce; in France rituximab concentrates in progressive disease. These results indicate that rituximab is an evidence-supported, cheaper option with room for wider use, not that any system should convert wholesale or that all lower utilization is unwarranted. An MS International Federation consensus similarly supports off-label rituximab for relapsing and active progressive MS where access to licensed drugs is constrained.[64]

The Italy and United Kingdom estimates rest on different footings. UK rituximab sits outside the NICE/Blueteq pathway that funds ocrelizumab and ofatumumab and is reachable only by Individual Funding Request, so its low share reflects commissioning structure rather than a counting gap. Italy is less certain: a COVID-era Italian cohort implies a rituximab share above our 3%. Because both measured corrections in this study (Germany, France) revised rituximab upward, we treat these figures, Italy especially, as conservative ceilings on cost: any undercount widens the transatlantic gap we report but means intra-European savings from wider rituximab use should be read as upper bounds, not firm estimates.

## Conclusion

Across six countries, variation in prescribing and expenditure aligns far more closely with regulatory approval status than with therapeutic effectiveness. Systems that restrict off-label rituximab pay many times more per patient annually for treatment a randomized controlled trial has now shown to be non-inferior, and Sweden delivers effective MS care at a modeled $76,000–$248,000 less per patient in drug acquisition cost over five years. These findings underscore a need for neurology treatment compendia, systematic off-label outcome tracking, and cost transparency at the point of prescribing. The pattern is likely to extend to any condition in which an off-patent biologic competes with a newer, patent-protected alternative, and may prove more consequential as international reference pricing reshapes pharmaceutical markets: such policies act on the price of the on-label agent, not on the regulatory asymmetry that determines which agent is used. Until that asymmetry is addressed, healthcare systems will continue to prescribe on the basis of regulatory approval status rather than comparative clinical evidence.

## Data Availability

All data supporting this study are drawn from published sources, national registry reports, and publicly available price schedules, each cited in the manuscript and supplementary appendix. No individual patient-level data were used. Simulation code and input parameters are described in the supplementary appendix; the Monte Carlo model used 10,000 iterations with seed 20260806 and is reproducible from the parameters reported.

