## Supplemental Material for "Regulation-Driven Variation in Utilization and Cost of B-Cell Depleting Therapies for Multiple Sclerosis: A Cross-National Study"

This appendix documents the derivation of every utilization and cost input, the regulatory frameworks referenced in the main text, the comparative-effectiveness evidence base, and the statistical methods. Where a figure could not be traced to a primary source, it is identified as an estimate and its basis is stated.

**S1. Utilization**

Rituximab utilization is expressed as a share of patients receiving B-cell depleting therapy (rituximab or ocrelizumab). Ofatumumab and ublituximab are excluded. This denominator is applied uniformly across all six countries.

| Country | RTX share | Data source | Provenance |
| --- | --- | --- | --- |
| Sweden | 96% | Swedish MS Registry [S19] | Registry-measured |
| United States | 5.3% | MarketScan commercial claims [3] | National claims |
| France | 18.7% | OFSEP national cohort [S22] | Registry-measured |
| Germany | 8.2% | German MS Registry 2024 [S21] | Registry-measured |
| Italy | 3.0% | No national source | Framework-derived |
| United Kingdom | 2.3% | No national source | Framework-derived |

Sweden. The registry reports rituximab in more than 7,400 of 12,738 patients on disease-modifying therapy, 58% to 63% of all such therapy, and describes it as completely dominant. The ocrelizumab share is inferred (S1.1).

United States. Rituximab is 5.3% of anti-CD20 initiations in MarketScan for 2018 to 2020, approximately 4% to 7%. Pooling with the Optum cohort gives 7.4%, carried as a sensitivity analysis (S1.2).

France. Rituximab is 18.7% of current anti-CD20 therapy, 1,431 of 7,646 patients, concentrated in progressive disease: 22.6% of primary progressive, 9.5% of secondary progressive and 1.2% of relapsing-remitting patients.

Germany. Among 8,733 treated patients ocrelizumab is 24.7% and rituximab 2.2% of all disease-modifying therapy, giving rituximab 8.2% of anti-CD20 use. Initiation data agree [60].

Italy. A cohort assembled during the COVID-19 pandemic [S23] implies a higher value but oversamples anti-CD20 agents. Maniscalco et al. [34] report rituximab in 5 of 3,025 first-line therapy starts, a share of all first-line starts rather than of anti-CD20 use; re-based it gives 5 of 11 patients accrued largely before ocrelizumab launched in Italy, and is not used here. The 3.0% value is framework-derived, off-label use being restricted by Law 648/96.

United Kingdom. Off-label rituximab is reachable only by Individual Funding Request, 71% of which were declined [27], and lies outside the NICE and Blueteq pathway funding ocrelizumab and ofatumumab; approximately 9,000 patients receive ocrelizumab [58]. The 2.3% value is a structural low estimate.

**S1.1 Sweden: derivation of the ocrelizumab share**

The analysis requires one figure for Sweden: the proportion of patients receiving B-cell depleting therapy who receive ocrelizumab rather than rituximab. No source publishes it, so it is estimated, bounded above, and tested across the range the registry permits.

He et al. [50] report 2 of 90 anti-CD20 treatment episodes as ocrelizumab, or 2.2%. That cohort pools the MSBase registry with the Swedish MS Registry and ends in 2017 and September 2019 respectively, before ocrelizumab was widely available, so the current share is likely higher; 4% is adopted for the primary analysis.

The 2024 annual report of the Swedish MS Registry [S19] bounds it. Of 12,738 patients receiving disease-modifying therapy at the end of 2023, 72.5%, or approximately 9,235, received high-efficacy therapy, which the registry defines as the anti-CD20 agents together with natalizumab, and more than 7,400 received rituximab. The residual of 1,235 to 1,835 patients is shared with ofatumumab and natalizumab, so ocrelizumab cannot exceed 13% to 20% of anti-CD20 use even if the entire residual is assigned to it. Alping et al. [18] report Swedish ocrelizumab uptake as too low to permit inclusion in comparative analyses.

Sweden's weighted annual cost was recalculated across the full range the registry data permit, on the drug-acquisition-only basis used throughout this analysis. Ratios in this table are deterministic point estimates; the main text reports the mean of the simulated ratio distribution, which is marginally higher.

| Assumed ocrelizumab share | Weighted annual cost | United States : Sweden cost ratio | Status |
| --- | --- | --- | --- |
| 2% | $2,667 | 19.7× | Within registry bounds |
| 4% | $3,014 | 17.4× | Primary analysis |
| 10% | $4,056 | 12.9× | Within registry bounds |
| 20% | $5,792 | 9.1× | Registry-implied ceiling |

*At the registry-implied ceiling, an ocrelizumab share the registry data themselves render implausible, Sweden's weighted cost remains 9.1-fold below that of the United States. The Swedish point estimate is sensitive to this assumption; the ratio is not.*

**S1.2 Estimation of the United States rituximab share of anti-CD20 use**

**S1.2.1 Rationale**

Three of the five European comparator countries have a national or near-national registry from which the rituximab share of anti-CD20 use can be read directly. The United States has no equivalent instrument. Anti-CD20 utilization in the United States must therefore be assembled from administrative claims databases and from single health systems, each of which observes a structurally different slice of the population. The resulting estimates differ by an order of magnitude; each source and the direction of its likely bias is given below.

**S1.2.2 Correction to the previously reported value**

An earlier draft reported a United States rituximab share of 1.4%. That figure is a denominator error and is withdrawn. It originates in Kern and Cepeda, Table 3, in which rituximab accounts for 28 of 1,994 first-line disease-modifying therapy initiations (1.4%). Re-based to the anti-CD20 denominator used throughout this analysis, the same table gives rituximab 28 of 148 first-line anti-CD20 initiations (18.9%), and the third-line table gives rituximab 2 of 25 (8.0%).

**S1.2.3 National sources**

Two studies report United States anti-CD20 utilization on a national sampling frame.

Henderson et al. analyzed 153,846 disease-modifying therapy initiation episodes in MarketScan Commercial Claims and Encounters among adults aged 18 to 64, 2001 to 2020. eTable 2 reports 5,290, 5,907 and 4,312 new use episodes in 2018, 2019 and 2020, with rituximab at 0.3%, 0.2% and 0.4% of each year's initiations and ocrelizumab at 4.7%, 5.1% and 5.8%. Ocrelizumab is absent before 2017 and appears at 4.3% in its approval year; ofatumumab appears at 1.3% in 2020 only. Rituximab never exceeds 0.4% of initiations in the twenty-year series.

Pooling the three years in which both anti-CD20 agents were established gives rituximab 44.9 and ocrelizumab 800.0 initiation episodes, a rituximab share of anti-CD20 initiations of 5.3%. Adding 2020 ofatumumab initiations to the denominator gives 5.0%.

Two independent sources of uncertainty attach to that figure. Binomial sampling error on 45 of 845 gives a 95% interval of 3.9% to 6.9%. Separately, because the source percentages are published to one decimal place, rounding alone gives a range of 4.4% to 6.2%. The two are comparable in width, so the estimate is reported as 5.3% (approximately 4% to 7%), with an outer envelope of 3.1% to 8.0% at the corners of the rounding range. A binomial interval quoted alone on counts reconstructed from rounded percentages would be overconfident.

eTable 4 tests indication specificity by repeating the analysis after excluding patients with malignancy or rheumatic disease at baseline, reducing the denominator to 144,277 episodes. Rituximab falls in every year of the anti-CD20 era, to 0.2%, 0.1% and 0.3%, while ocrelizumab is unchanged. Pooled and re-based, the rituximab share falls from 5.3% to 3.5% (rounding envelope 2.6% to 4.4%), so some rituximab initiations in the unrestricted table likely reflect haematologic or rheumatologic indications. The restriction may over-correct, since multiple sclerosis co-occurs with both comorbidities, so 3.5% is carried as the indication-restricted lower sensitivity rather than as the primary value.

Kern and Cepeda analyzed Optum Clinformatics, 5,691 patients with incident multiple sclerosis between 2014 and 2019. Combining the reported first-line and third-line anti-CD20 initiations gives rituximab 30 of 173 (17.3%, 95% CI 12.3% to 23.5%). This source is not commercial-only: Table 1 of that paper reports 41.2% of the cohort enrolled in Medicare Advantage plans, with the remainder commercially insured.

Geiger et al. (Optum Market Clarity, n = 682) provide an upper bound only. Patients treated with rituximab alone were excluded by design, fewer than 33 and under 5% of the eligible sample, and rituximab appears in the results only within an "other disease-modifying therapy" category at fewer than 10 patients against 113 for ocrelizumab, implying a share below 8%.

**S1.2.4 Composite national estimate**

Pooling the two national cohorts on raw counts gives 75 of 1,018, or 7.4% (95% CI 5.9% to 9.1%). The two series are, however, highly heterogeneous (Cochran Q = 27.2 on 1 degree of freedom, I² = 96%), so a random-effects pool on the logit scale returns 9.7% with a 95% interval of 2.9% to 28.2%, which is too wide to be informative.

The heterogeneity is structural rather than sampling error. Kern and Cepeda accrued incident patients from 2014, so more than half the cohort predates ocrelizumab's 2017 approval and its anti-CD20 denominator is depleted of ocrelizumab; Henderson et al. observed 2018 to 2020, after ocrelizumab was established, and is the larger and more recent series. Payer mix does not account for the gap, since the cohort with the higher rituximab share is the one including Medicare Advantage enrollees. The accrual window alone explains the divergence.

We adopt Henderson et al.'s 5.3% as the primary United States value, carry the 7.4% pooled figure as a sensitivity analysis, and treat 17.3% as the upper limit of the national claims evidence.

**S1.2.5 Regional and health-system variation**

Estimates from individual United States health systems diverge from the national claims figures and are reported as evidence of variation, not folded into the national estimate.

Kwon et al. found rituximab in 922 and ocrelizumab in 956 patients in the University of Colorado Health System linked to the Colorado All Payer Claims Database, 2018 to 2020, a rituximab share of 49.1%. Its authors attribute this to longstanding rituximab use at the reporting center and easy access through Colorado Medicaid. Counts are unique users per epoch, so patients who switched are counted in both drugs, which biases the reported share downward.

Langer-Gould et al. reported approximately 80% of patients on highly effective therapy at Kaiser Permanente Southern California in 2018 receiving rituximab. Neither ocrelizumab nor ofatumumab was available in that system during the study period, so its anti-CD20 share is complete by construction and the cohort is informative about system-level standardization rather than about a choice between the two agents.

Kwon et al. attribute their divergence from commercial claims cohorts to capture of infusions from the medical claim line and to all-payer rather than commercial-only coverage, citing Henderson et al. as the comparator showing low infusion adoption. The within-country spread reflects data-source structure and local practice, not clinical disagreement.

**S1.2.6 Independent cross-check from the MSIF Atlas of MS**

The MSIF Atlas of MS, Part 2, reports the proportion of people treated with each disease-modifying therapy, a utilization measure independent of United States claims data. Rituximab and ocrelizumab account for 5% and 8% of people on therapy globally, 3% and 10% in the WHO European Region, and 2% and 9% in high-income countries. Re-based onto the anti-CD20 denominator these are rituximab shares of approximately 39%, 23% and 18%.

Only 57 countries supplied proportions, values are published rounded to whole numbers (so the high-income figure spans 14% to 23%), and MSIF notes wide variation within income bands and WHO regions. They are reported only as an independent indication that the rituximab share of anti-CD20 use in high-income settings lies in the low tens of percent.

The WHO European Region figure of 23% is close to the unweighted mean of the five European shares reported here (26%) rather than their population-weighted mean (11%), as expected when the Atlas aggregates by country and registries by patient. That region includes Türkiye, Israel, the Russian Federation and the Central Asian republics and is neither the European Union nor geographic Europe.

The Atlas supplies no United States figure: with the United States set as country of interest, every disease-modifying therapy row of the utilization table returns “used, but unsure of percentage”.

**S1.2.7 Unmeasured strata**

The primary estimate rests on commercially insured adults aged 18 to 64, approximately 55% of the covered United States population. Medicare Advantage enrollees are partly observed through the Kern and Cepeda cohort, but no study reports a rituximab share in fee-for-service Medicare, Medicaid, Veterans Affairs or uninsured populations. Kim et al. report national ocrelizumab volumes in Medicare (9,924 treatment courses) and Medicaid (4,972) for 2021 but cannot attribute rituximab volume to multiple sclerosis, since it appears in both datasets without indication. Roughly half the prevalent population is therefore unobserved for this parameter.

To test how much the unobserved strata could matter, a population-weighted Bayesian random-effects synthesis was fitted, with the unobserved remainder imputed from the estimated between-system distribution. It returns a national share of 19.8% (95% credible interval 9.1% to 34.3%), but that value moves between 11% and 49% according to which measured cohort carries weight, and its entire distance from Henderson's 5.3% is generated by the imputation rather than by observation. The synthesis therefore bounds the fragility of extrapolating to the unobserved half of the population rather than estimating it, and the measured value is retained as primary.

**S1.2.8 Absolute magnitude**

Scaling Kim et al.'s public-payer ocrelizumab volumes to the national level on the assumption that commercial insurance covers half of treated patients implies roughly 30,000 patients receiving ocrelizumab for multiple sclerosis in the United States. At a 5.3% rituximab share this corresponds to approximately 1,700 patients receiving rituximab for multiple sclerosis nationally, and at the 7.4% pooled share to approximately 2,400. At the indication-restricted 3.5% share the figure falls to approximately 1,100. In every case this is under 0.3% of the estimated 913,925 prevalent United States multiple sclerosis population. Rituximab use for multiple sclerosis in the United States is therefore numerically negligible outside individual health systems, against a large majority of anti-CD20 use in Sweden.

**S1.2.9 The initiation frame**

Every United States estimate above is a share of initiations, whereas the cost model prices prevalent patients. Henderson et al. count an episode as new use when there has been no claim for that agent in the preceding 365 days, so patients switched onto rituximab do enter the numerator and 26.1% of adults contributed more than one episode. What the frame cannot capture is a patient maintained on rituximab continuously from before the window. Because ocrelizumab initiations in 2018 to 2020 are almost entirely new starts by construction, the frame understates a stable off-label incumbent relative to a recently launched brand.

No published source closes the gap. Henderson's prevalent-use analysis (eFigure 1) plots only frequently used therapies and omits rituximab, and the persistence literature yields no usable multiplier: the one United States cohort with the appropriate design excluded off-label agents by protocol, and the two non-United States cohorts disagree in sign. The direction is therefore reported rather than a corrected value, the national figures understating the prevalent share by an amount the record does not identify. Kwon et al.'s Colorado cohort is the only United States measurement on a prevalent basis (S1.2.5). The understatement is bounded, since there is no large earlier rituximab cohort for the washout to conceal (S1.2.3). Reconstructing prevalent stock from the published initiation series under annual rituximab discontinuation rates of 5% to 30%, against the reported 13% for ocrelizumab, places the prevalent share between roughly 5% and 13%, mid-range near 7%. The pooled 7.4% estimate therefore lies inside that range, and the United States to Sweden ratio varies only between 16.3 and 17.7 across the whole of it, including the indication-restricted lower bound.

**S1.2.10 Direction of bias**

A lower assumed rituximab share raises the estimated United States weighted average annual cost and therefore widens the apparent gap with the European comparators. The low end of the range is the direction favorable to the argument advanced here. Across the full combined interval the United States weighted average annual cost moves from $53,186 per patient-year at a 3.9% share, through $52,506 at 5.3% and $51,486 at the pooled 7.4%, to $46,676 at 17.3%. The corresponding United States to Sweden point-estimate ratios are 17.6, 17.4, 17.1 and 15.5.

Two biases act in opposite directions and partly offset. Restricting to indication-specific initiations lowers the share to 3.5%, raising the weighted average to $53,392 and the ratio to 17.7; moving to the prevalent frame raises the share by an unidentified amount. Only the first is quantified, so the unrestricted measured value is primary and both departures are reported as named directions rather than a net adjustment.

The central quantitative claim of this analysis does not depend on the United States rituximab share. Because United States and Swedish ex-factory prices differ by the same factor for both agents (rituximab $6,500 against $2,320, and ocrelizumab $55,081 against $19,678, both 2.80-fold), the United States to Sweden cost ratio separates into a price component and a utilization component. Evaluated at Sweden's own utilization mix, the price component is 2.80-fold ($8,443 against $3,014 per patient-year for an identical therapeutic strategy), and this quantity requires no estimate of United States utilization.

**S2. Pricing**

Annual drug costs by country. Derivation and the treatment of confidential discounts follow the table.

| Country | RTX/yr | OCR/yr | Primary source |
| --- | --- | --- | --- |
| Sweden | $2,320 | $19,678 | Alping et al. [18] |
| France | $747 | $22,140 | Journal Officiel [S15, S16] |
| United Kingdom | $2,218 | $24,333 | NICE TA533 [26]; NHS England [63] |
| Germany | $3,032 | $22,144 | Lauer-Taxe / Gelbe Liste Pharmindex [S18] |
| Italy | $2,398 | $27,000 | Gazzetta Ufficiale, AIFA [61, 62] |
| United States | $6,500 | $55,081 | ICER Final Evidence Report 2023 [31, 32] |

**S2.1 Derivation by country**

Sweden. Alping et al. [18] assign rituximab $1,160 per 500 mg vial plus $400 per infusion visit; at 500 mg every 6 months this is $3,120 per year including administration. The table and model use the drug-acquisition component of $2,320 only, so Sweden sits on the same basis as the other five countries. Ocrelizumab is assigned $9,839 per 600 mg dose every 6 months, or $19,678 per year. These are reference unit costs in that cost model, not observed procurement prices.

France. Both figures are read from the Journal Officiel at a matched date, in force 1 March 2024. Ocrelizumab carries a tarif de responsabilité of €5,124.998 per 300 mg vial (UCD 3400894351368) at 4 vials per year [S15]. Rituximab carries a tarif unifié of €345.929 per 500 mg vial at 2 vials per year [S16], applied identically to the originator and to all biosimilars.

United Kingdom. NICE TA533 states a list price of £4,790 per 300 mg ocrelizumab vial; at 4 vials per year this is £19,160 ($24,333) [26]. The NHS England Integrated Impact Assessment states £873.15 per 500 mg/50 mL MabThera vial excluding VAT; at 2 vials per year this is £1,746 ($2,218) [63]. NHS England notes rituximab is negotiated under local arrangements, so the real acquisition cost is below list.

Germany. Pharmacy retail prices are read from the Lauer-Taxe [S18]: ocrelizumab €6,340.14 per 300 mg vial (PZN 11688732) and €12,621.85 for the two-vial pack (PZN 15744002), rituximab €1,778.11 (Ituxredi 500 mg, PZN 19074515). That price includes 19% value-added tax and pharmacy and wholesale margins and is not what hospitals pay. German databases do not publish the manufacturer price, so it is derived by reversing the Arzneimittelpreisverordnung, giving €5,125.86 per 300 mg ocrelizumab vial (€20,503 per year) and €1,403.88 per 500 mg rituximab vial (€2,808 per year). The derivation is validated twice: the single-vial and two-vial pack sizes independently yield €5,125.86 and €5,125.43 despite different fixed-fee structures, and the result matches the French government tariff for the identical vial (€5,124.998) to within €0.86.

Italy. Both figures are read from the Gazzetta Ufficiale. Ocrelizumab has an ex-factory price, excluding value-added tax, of €6,250.00 per 300 mg vial in class H (GU Serie Generale n. 204, 3 September 2018, reconfirmed by Determina of 21 February 2022), giving €25,000 per year at 4 vials [61]. The Ruxience rituximab biosimilar (A.I.C. 048720027) has an ex-factory price of €1,110.17 per 500 mg vial (GU n. 45, 24 February 2021), giving €2,220 per year at 2 vials [62]. An earlier draft used €9,309.29 per vial, a retail-basis figure; ocrelizumab is class H and hospital-dispensed, so the ex-factory price applies.

United States. Both figures are read from the ICER Final Evidence Report 2023 [31, 32]. Ocrelizumab at $55,081 is a manufacturer-provided net price, a 23% discount to the $71,187 wholesale acquisition cost; the 6% provider-administered mark-up is excluded here as in that report. ICER gives rituximab as $4,000 to $9,000 annually, biosimilar average sales price approximately $4,400; the midpoint is used.

**S2.2 Pricing basis**

All prices are reported on a consistent ex-factory basis, the manufacturer price excluding value-added tax and distribution margins. These agents are hospital-administered and only exceptionally dispensed through community pharmacies, and value-added tax on medicines differs between the countries studied (Germany 19%, Italy 10%, France 2.1%), so tax-inclusive comparison would conflate fiscal policy with drug pricing. Ex-factory is the basis used by the OECD, WHO and EURIPID for cross-national comparison.

Every price in this table is a list price. Beneath each sits a confidential discount that no public source discloses: Germany’s AMNOG-negotiated Erstattungsbetrag, the UK patient access scheme, Italy’s sconto obbligatorio, and French hospital tenders. Italian regions additionally tender rituximab biosimilars below the published ceiling. No discount percentage has been assumed or imputed anywhere in this analysis.

A further consideration applies to Germany. Ocrelizumab may be purchased through hospital channels at or below the manufacturer price, or dispensed through a community pharmacy at the retail price in office-based practice, in which case total system outlay exceeds the price reported here. The manufacturer price is used because it is the quantity this study concerns, because the comparator countries purchase through hospital channels, and because Germany’s AMNOG reimbursement amount is negotiated at that level.

The United States is the single exception, since ICER publishes an explicit net-of-rebate figure, which is used here. Comparing European list prices against a United States net price is conservative with respect to the central claim of this analysis: the United States wholesale acquisition cost is $71,187, and using it would widen the transatlantic difference rather than narrow it.

Dosing assumptions are corroborated by the OVERLORD-MS randomized trial [51], which administered rituximab as 1,000 mg at baseline then 500 mg every 6 months (maintenance 1,000 mg/year) and ocrelizumab as 600 mg every 6 months (1,200 mg/year), matching the regimens used throughout.

All costs are expressed in 2024 US dollars. Euro figures are converted at EUR/USD 1.08 and sterling at GBP/USD 1.27.

*ICER additionally reports that the price premium between rituximab and ocrelizumab's net price in the United States is between 600% and 1,300% [31].*

**S3. Regulatory Frameworks**

Germany, the Sandoglobulin decision. BSG B 1 KR 37/00 R (2002) [25] established that statutory health insurance must fund off-label treatment only where three conditions are met: a life-threatening or seriously debilitating illness; no available approved alternative; and a realistic prospect of benefit. In MS, the availability of multiple approved disease-modifying therapies means the second condition is generally unmet, and off-label rituximab is consequently not routinely reimbursed. Physicians additionally face liability exposure under the Arzneimittelgesetz [S8].

United Kingdom, Individual Funding Requests. Off-label rituximab for MS requires an Individual Funding Request to NHS England [28]. Approval requires demonstration of clinical "exceptionality", that the patient differs materially from others with the same condition. Sanghvi et al. [27] found 71% of IFRs to NHS England were declined, most commonly for failure to demonstrate exceptionality (42/45; 93% of declines).

Italy, Law 648/96. Law 648/96 [35] permits reimbursement of drugs for unapproved indications when no valid therapeutic alternative exists. Approved disease-modifying therapies exist, so rituximab does not qualify, and it is not included on the AIFA 648 list for MS.

**S4. Comparative Effectiveness Evidence**

| Study | Ref | Findings | Design |
| --- | --- | --- | --- |
| OVERLORD-MS | [51] | Phase 3, double-blind, noninferiority RCT. 216 treated (132 RTX / 84 OCR). Rituximab met noninferiority on the primary MRI endpoint (92.2% vs 94.8% free of new/enlarging T2 lesions; risk difference −2.6 pp, 95% CI −9.4 to 4.3; P=0.03). ARR 0.09 vs 0.04. Six-month confirmed disability progression 3% vs 7%. | Randomized |
| RIFUND-MS | [17] | Phase 3 RCT, 200 patients randomized 1:1 across 17 Swedish hospitals. Rituximab reduced relative relapse risk by 81% versus dimethyl fumarate (3% [3/98] vs 16% [16/97] relapsing over 24 months; risk ratio 0.19, 95% CI 0.06–0.62). | Randomized |
| Roos et al. | [7] | MSBase and Danish MS Registry. From 1,613 patients, propensity matching yielded 710 ocrelizumab- and 186 rituximab-treated. Rituximab did not meet noninferiority for relapse control (ARR 0.20 vs 0.09; rate ratio 1.8, 95% CI 1.4–2.4). Confirmed disability accumulation did not differ. | Observational |
| Granqvist et al. | [19] | Swedish population-based cohort, 494 patients initiating first DMT. Rituximab annual discontinuation 0.03 vs 0.53 (injectables), 0.32 (dimethyl fumarate), 0.38 (fingolimod), 0.29 (natalizumab); lower relapse and radiological activity than injectables and dimethyl fumarate. | Observational |
| Savolainen et al. | [52] | Finnish population-based matched cohort, 2018–2024. 636 patients screened, 191 eligible, 112 matched (56 rituximab, 56 ocrelizumab). ARR 0.03 in both arms; relapse-free survival did not differ. | Observational |
| Salzer et al. | [16] | 822 rituximab-treated patients; annualized relapse rate 0.044 in RRMS over a mean 21.8 months' follow-up. | Observational |
| Virtanen et al. | [20] | Nationwide Swedish cohort, 4,694 rituximab-treated RRMS patients (2012–2021). Serious infection rate approximately double that of comparator DMTs (22.7 vs 10.4 per 1,000 person-years), but risk did not vary by treatment line, prior therapy, or exposure time. | Observational |
| ROC-MS | [36] | Prospective individual-participant-data meta-analysis pooling four randomized trials (1,109 patients; 660 rituximab, 449 ocrelizumab). Results expected 2028. | Pending |

*Beyond MS, rituximab is used off-label in neuromyelitis optica spectrum disorder [49] and myasthenia gravis [48], where the same regulatory asymmetry between an off-patent agent and its patent-protected successors applies.*

**S5. Statistical Methods**

Monte Carlo simulation (10,000 iterations) was used to estimate cost distributions and 95% credible intervals. Weighted average annual cost per patient receiving B-cell depleting therapy was calculated as:

*Cost = (RTX share × RTX price) + (OCR share × OCR price)*

Price inputs were varied about their point estimates; utilization inputs were drawn from beta distributions for countries with estimated shares and held fixed for countries with registry-derived or model-derived shares.

**S5.1 Savings estimation**

Per-patient five-year savings ($35,000–$66,000) derive directly from Alping et al. [18], which compared total healthcare costs for off-label rituximab against approved DMTs in 5,924 Swedish therapy starts. This figure is reported verbatim in the source and is the primary savings result.

Country MS populations are taken from the Atlas of MS (3rd edition, 2020) [21] on a single consistent basis: United States 913,925; Germany 280,000; United Kingdom 133,780; Italy 133,000; France 100,000; Sweden approximately 21,900 (total ≈ 1.58 million).

Per-patient savings apply only to patients receiving B-cell depleting therapy, so the aggregate is restricted to that subset. Applying a B-cell-therapy treatment rate of approximately 5–7% (consistent with US utilization modeled by ICER, where roughly 50,000 of ≈900,000 MS patients received B-cell depleting therapy) gives an exposed population of 80,000–110,000 patients across the six countries, and an exploratory aggregate five-year saving on the order of $3–$7 billion on the total healthcare cost basis of Alping et al. This is a different quantity from the $13–$18 billion reported in the main text, which applies the same exposed population to the modeled drug-acquisition cost differential of this study (a population-weighted $165,700 per patient over five years) rather than to Alping's empirical $35,000–$66,000. Both are scenario illustrations, not projections: treatment-eligible population sizes are not uniformly reported across countries.

**S6. Additional Limitations**

Utilization figures of national scope now exist for Sweden, Germany, and France, from national registries; the United States is measured from national commercial claims (S1.2 [3]). Italy and the United Kingdom remain framework-derived estimates. Measured initiation data corroborate the underlying premise for the UK (rituximab 0.1% of first DMT starts [60]), but these two countries do not publish a prevalence share of B-cell depleting therapy; Italy is the least supported. Adopting a higher rituximab share for either country would reduce its weighted cost and narrow, rather than widen, the gap reported here.

European prices are published list prices while the US figure is net of rebates; confidential discounts are not public. Indirect costs, including productivity loss and caregiver burden, were not assessed. Our claims are correspondingly associational rather than causal.

**Supplementary References**

S1. Tandvårds- och läkemedelsförmånsverket (TLV). Hälsoekonomisk bedömning av Ocrevus vid behandling av multipel skleros. Stockholm: TLV.

S2. Ahlgren C, Odén A, Lycke J. High nationwide prevalence of multiple sclerosis in Sweden. Mult Scler. 2011;17(8):901-908.

S3. Federal Statistical Office of Germany (Destatis). Health reporting data on multiple sclerosis. Wiesbaden: Destatis.

S4. Public Health England. Multiple sclerosis: prevalence, incidence and smoking status — data briefing. London: PHE; 4 February 2020.

S5. Multiple Sclerosis International Federation. Atlas of MS, 3rd edition (2020). London: MSIF; 2020. https://www.atlasofms.org

S6. Battaglia MA, Bezzini D. Estimated prevalence of multiple sclerosis in Italy in 2015. Neurol Sci. 2017;38(3):473-479.

S7. Papukchieva S, Stratil AS, Kahn J, et al. Shifting from treat-to-target to early highly effective treatment in multiple sclerosis. Ther Adv Neurol Disord. 2022;15:17562864221118729.

S8. German Medicines Act (Arzneimittelgesetz, AMG). Sections 96–97: criminal liability provisions relating to off-label prescribing.

S9. Spelman T, Magyari M, Piehl F, et al. Treatment escalation vs immediate initiation of highly effective treatment for patients with relapsing-remitting multiple sclerosis. JAMA Neurol. 2021;78(10):1197-1204.

S10. Vesperinas-Castro A, Cortés-Vicente E. Rituximab treatment in myasthenia gravis. Front Neurol. 2023;14:1275533. (See main reference [48].)

S11. Hayes MTG, Adam RJ, McCombe PA, Walsh M, Blum S. Long-term efficacy and safety of rituximab in the treatment of neuromyelitis optica spectrum disorder. (See main reference [49].)

S12. Gemeinsamer Bundesausschuss (G-BA). Beschluss über eine Änderung der Arzneimittel-Richtlinie: Anlage XII — Ocrelizumab. 2 August 2018.

S13. Cortesi PA, Paolicelli D, Capobianco M, et al. The value and sustainability of ocrelizumab in relapsing multiple sclerosis: a cost-effectiveness and budget impact analysis. Farmeconomia Health Econ Ther Pathways. 2019;20(1):61-72.

S14. Moccia M, Affinito G, Berera G, et al. Persistence, adherence, healthcare resource utilization and costs for ocrelizumab in the real world of the Campania Region of Italy. J Neurol. 2022;269(12):6504-6511.

S15. Avis relatif aux prix de spécialités pharmaceutiques publiés en application de l'article L. 162-16-6 du code de la sécurité sociale (ocrelizumab). NOR TSSS2402521V. JORF n°0024 du 30 janvier 2024, texte n°90. Tarif en vigueur au 1er mars 2024.

S16. Avis relatif aux prix de spécialités pharmaceutiques (rituximab, tarif unifié), art. L. 162-16-6 du code de la sécurité sociale. NOR TSSS2403781V. JORF n°0033 du 9 février 2024, texte n°107. Tarif en vigueur au 1er mars 2024.

S17. Agenzia Italiana del Farmaco (AIFA). Liste dei farmaci di classe H — elenco dei medicinali e relativi prezzi. Ocrelizumab (Ocrevus 300 mg), AIC 045889019. Rome: AIFA. Accessed 16 June 2026.

S18. Gelbe Liste Pharmindex (ABDATA / Lauer-Taxe). Arzneimittelpreisverzeichnis. Ocrevus 300 mg Konzentrat: PZN 11688732 (1 vial, AVP €6,340.14) and PZN 15744002 (2-vial N1 pack, AVP €12,621.85); Ituxredi 500 mg: PZN 19074515. https://www.gelbe-liste.de — accessed 1 July 2026.

S19. Svenska MS-registret. Årsrapport för 2024: Multipel skleros [Swedish MS Registry Annual Report 2024]. Stockholm: Svenska Neuroregister; 2025 (revised 11 August 2025). https://www.neuroreg.se/media/bryfepkb/ms-årsrapport-2024-justerad-2025-08-11.pdf

S20. Stahmann A, Craig E, Ellenberger D, et al. Disease-modifying therapy initiation patterns in multiple sclerosis in three large MS populations. Ther Adv Neurol Disord. 2024;17:17562864241233044. (See main reference [60].)

S21. Multiple Sklerose Register der DMSG, Bundesverband e.V. Berichtsband 2024. Hannover: MS Forschungs- und Projektentwicklungs-gGmbH; 2025:32 (Abbildung 27, Wirkstoffverteilung der aktuellen immunmodulatorischen Therapie). https://www.msregister.de

S22. Observatoire Français de la Sclérose en Plaques (OFSEP). Descriptif de la cohorte — traitement de fond en cours des patients en file active (export 8 décembre 2025). https://www.ofsep.org/fr/la-cohorte-ofsep/descriptif-de-la-cohorte

S23. Sormani MP, Salvetti M, Labauge P, et al. DMTs and Covid-19 severity in MS: a pooled analysis from Italy and France. Ann Clin Transl Neurol. 2021;8(8):1738–1744. doi:10.1002/acn3.51408

**S7. CHEERS 2022 checklist**

Consolidated Health Economic Evaluation Reporting Standards 2022, completed for this manuscript. This study is a cross-national cost and utilization analysis rather than a full economic evaluation: it compares drug-acquisition cost per treated patient between health systems and does not estimate incremental cost per unit of health gain. Items concerning discounting, outcome measurement and valuation, and distributional modelling (items 10 to 13, 19, 21 and 25) are therefore reported as not applicable with the reason stated, rather than left blank.

| Section | No. | CHEERS 2022 item | Reported in / response |
| --- | --- | --- | --- |
| Title | 1 | Title | Title page. Identifies the study as a cross-national analysis of utilization and cost; the two interventions compared are named in the abstract and throughout. |
| Abstract | 2 | Abstract | Abstract, structured under Background, Objective, Methods, Results and Conclusion. |
| Introduction | 3 | Background and objectives | Introduction. States the regulatory asymmetry between an off-patent off-label agent and a licensed patent-protected successor, and the policy question. |
| Methods | 4 | Health economic analysis plan | No prospective plan was registered. The full analytic specification, including every input and its provenance, is given in S1, S2 and S5. |
| Methods | 5 | Study population | Methods and S1. The unit of analysis is a patient receiving anti-CD20 therapy for multiple sclerosis in each system; national populations and registry coverage are in Table 2 and S5.1. Individual-level data were not used. |
| Methods | 6 | Setting and location | Methods, Results, Table 2. Six health systems, 2016 to 2024. Regulatory frameworks are described in Results and S3. |
| Methods | 7 | Comparators | Introduction and Methods. Rituximab and ocrelizumab target the same antigen and differ in regulatory status, isolating regulation from mechanism. Ofatumumab and ublituximab are excluded from the denominator. |
| Methods | 8 | Perspective | Methods. Payer perspective, restricted to drug acquisition cost on a consistent ex-factory basis. Administration, monitoring and indirect costs are excluded, as stated in Limitations. |
| Methods | 9 | Time horizon | Methods and Results. Primary results are annual cost per treated patient; a five-year horizon is used for the illustrative differences, matching the Swedish cohort study from which the empirical savings figure is taken. |
| Methods | 10 | Discount rate | Not applicable. Costs are not discounted; the five-year figures are undiscounted sums of a constant annual difference and are presented as illustrative differences rather than present values. |
| Methods | 11 | Selection of outcomes | Not applicable. This is a cost and utilization analysis, not a cost-effectiveness analysis; no health outcome is used as a denominator. Comparative effectiveness and safety are summarized in the Discussion and tabulated in S4. |
| Methods | 12 | Measurement of outcomes | Not applicable to the economic model. Clinical endpoints in the Discussion are taken as published; designs and effect estimates are in S4. |
| Methods | 13 | Valuation of outcomes | Not applicable. No health-state valuation, utility weighting or quality-adjusted life-year estimation was undertaken. |
| Methods | 14 | Measurement and valuation of resources and costs | Methods and S2, S2.1, S2.2. Every price is read from a primary national source on an ex-factory basis; the German manufacturer price is derived by reversing the statutory formula and validated twice. Confidential discounts are not imputed. |
| Methods | 15 | Currency, price date, and conversion | Methods and S2. 2024 US dollars; EUR/USD 1.08, GBP/USD 1.27. Price dates are given per country in S2.1. |
| Methods | 16 | Rationale and description of model | Methods and S5. A weighted-average cost model is used because no source reports cost per treated patient directly. The model is fully specified by Table 1; code is available from the corresponding author. |
| Methods | 17 | Analytics and assumptions | Methods and S1.1, S1.2, S5. Utilization shares are re-based onto a common anti-CD20 denominator; the derivation of the Swedish and United States shares, including sources rejected and why, is in S1.1 and S1.2. |
| Methods | 18 | Characterizing heterogeneity | Results, Limitations, S1.2.5. Within-country heterogeneity is reported rather than pooled: United States estimates range from 5.3% nationally to 49.1% in one all-payer state cohort. Variation by phenotype is reported for France. |
| Methods | 19 | Characterizing distributional effects | Not formally modelled. Equity-relevant coverage gaps are reported descriptively: the United States estimate observes commercially insured adults aged 18 to 64 only, leaving fee-for-service Medicare, Medicaid, Veterans Affairs and uninsured populations unmeasured (S1.2.7). |
| Methods | 20 | Characterizing uncertainty | Methods and S1.1, S1.2, S5. Monte Carlo simulation with 10,000 iterations propagates price uncertainty for all six countries and utilization uncertainty for the two framework-derived shares. Deterministic sensitivity analyses span both contested shares. No significance tests are applied to simulated output, and the reason is stated. |
| Methods | 21 | Engagement with patients and others affected | No patient, public or stakeholder engagement was undertaken. The analysis uses published aggregate data only. |
| Results | 22 | Study parameters | Table 1 reports every price and utilization input with its source; S1 and S2 give provenance, derivation and limitations; S5 gives distributional assumptions. |
| Results | 23 | Summary of main results | Results, Table 1, Figure 2. Mean annual cost per treated patient with 95% credible intervals for each country, and cost ratios relative to Sweden. |
| Results | 24 | Effect of uncertainty | Results, Limitations, S1.1 and S1.2.10. Credible intervals throughout. Sensitivity of the headline ratio is quantified across the full plausible range of both contested shares; the United States to Sweden ratio remains between 16.3 and 17.7. Discounting is not applied. |
| Results | 25 | Effect of engagement with patients and others affected | Not applicable; no such engagement was undertaken. |
| Discussion | 26 | Study findings, limitations, generalizability | Discussion and Limitations, with additional limitations in S6. Covers the list-price versus net-price asymmetry, the framework-derived shares for Italy and the United Kingdom, the initiation versus prevalence frame for the United States, phenotype mixing, and excluded cost categories. |
| Other | 27 | Source of funding | Title page, Funding statement. To be completed by the authors. |
| Other | 28 | Conflicts of interest | Title page, Conflict of Interest statement, with ICMJE forms. To be completed by the authors. |
